# External Control Arm with Synthetic Real-world Data for Comparative Oncology using Single Trial Arm Evidence (ECLIPSE): A Case Study using Lung-MAP S1400I

**DOI:** 10.1101/2024.09.10.24313417

**Authors:** Alind Gupta, Luke Segars, David Singletary, Johan Liseth Hansen, Kirk Geale, Anmol Arora, Manuel Gomes, Ramagopalan Sreeram, Winson Cheung, Paul Arora

**Affiliations:** Inka Health, Canada; Division of Epidemiology, Dalla Lana School of Public Health, University of Toronto, Canada; Subsalt, North Carolina, USA; Quantify Research, Stockholm, Sweden; University of Cambridge, London, UK; University College London Hospital, London, UK; University College London, London, UK; Centre for Pharmaceutical Medicine Research, King’s College London, London, UK; Department of Oncology, University of Calgary, Alberta, Canada

## Abstract

2.

Single-arm trials supplemented with external comparator arm(s) (ECA) derived from real-world data are sometimes used when randomized trials are infeasible. However, due to data sharing restrictions, privacy/security concerns, or for logistical reasons, patient-level real-world data may not be available to researchers for analysis. Instead, it may be possible to use generative models to construct synthetic data from the real-world dataset that can then be freely shared with researchers. Although the use of generative models and synthetic data is gaining prominence, the extent to which a synthetic data ECA can replace original data while preserving patient privacy in small samples is unclear.

**Objective:** To compare the efficacy of nivolumab + ipilimumab combination therapy (“experimental arm”) versus nivolumab monotherapy (“control arm”) in patients with metastatic non-small cell lung cancer (mNSCLC) using real-world data from two real-world databases (“original ECA”), and synthetic data versions of these datasets (“synthetic ECA”), with the aim of validating synthetic data for use in ECA analysis.

**Study design:** Non-randomized analyses of treatment efficacy comparing the experimental arm to the (i) original ECA and (ii) synthetic ECA, with baseline confounding adjustment.

**Data sources:** The experimental arm is from the Lung-MAP no-match substudy S1400I (<u>NCT02785952</u>) provided by National Clinical Trials Network (NCTN) in the United States. The real-world data source for the ECA is data from population-based oncology data from the Canadian province of Alberta, and from Nordic countries in Europe, specifically Denmark and Norway.

## 3. Amendments and updates

| Version date | Version number | Section of protocol | Amendment or update | Reason |
| --- | --- | --- | --- | --- |
| March 12, 2024 | 0.1 | All | First draft | n/a |
| July 23, 2024 | 0.2 | All | Update | Including additional data source, and expanding sections on benchmarking metrics and specific generative models; Update timelines |
| September 3 | 0.3->1.0 | All | Update | Revisions based on feedback from AA and JLH on version 0.2 of this document |

## 4. Milestones

**Table 1.** Milestones.

| Milestone | Date |
| --- | --- |
| Project initiation<br>• Finalize study objectives and scope of work<br>• Establish tasks and timelines<br>• Provide communication channels | September 2024 |
| Data preparation | October 2024 |
| Statistical analysis including synthetic data generation | December 2024 |
| Discussion of results with all collaborators | February 2025 |
| Manuscript draft and revisions | March 2025 |
| Submission to scientific journal | April 2025 |

## 5. Rationale and background

Clinical researchers often encounter challenges in accessing patient-level real-world data for external comparator arms (ECAs) in single-arm trials, attributed to data sharing restrictions, privacy concerns, and logistical constraints. To address this issue, an emerging approach involves utilizing generative models to construct synthetic data that mirrors the statistical properties of real-world datasets. This strategy enables the creation of alternative comparator arms, which can be freely shared and analyzed, circumventing obstacles associated with data availability. However, the extent to which synthetic data faithfully reproduces the characteristics of the original real-world dataset remains uncertain, raising questions about its reliability and validity in the context of single-arm trials.

This research aims to explore the viability of synthetic data in external comparator arms by assessing its fidelity compared to real-world data. The study delves into the current landscape of single-arm trials, emphasizing the limitations in accessing patient-level real-world data and elucidating the role of synthetic data in overcoming these challenges. By investigating the potential benefits and limitations of synthetic data generation methods, the research seeks to provide valuable insights into the reliability of study outcomes derived from ECAs constructed using generative models, thereby contributing to the ongoing discourse on innovative methodologies for enhancing the robustness of single-arm trials amidst data constraints.

The following are details about the Lung-MAP trial used in this study:

**What is known about the condition:** Squamous cell lung carcinoma is a histological subtype of NSCLC that originates in the squamous cells lining the airways of the lungs. Historically, 25-30% of all cases of NSCLC are squamous cell carcinoma although these percentages can vary regionally and may change over time due to factors such as changes in smoking patterns. Compared to other NSCLC subtypes, such as adenocarcinoma, the presence of actionable genetic variants is less common and there are fewer targeted therapies available for squamous cell advanced/metastatic NSCLC.

**What is known about the exposure of interest:** Patients diagnosed with squamous mNSCLC often receive Platinum-based chemotherapy regimens as front-line systemic therapy. Following progression, patients will most often receive therapy with immune checkpoint inhibitors that target either the PD-(L)1 pathway, such as nivolumab and atezolizumab, or CTLA-4, such as ipilimumab. The Lung-MAP S1400I trial (NCT02785952) compared overall survival in United States patients with recurrent stage IV squamous NSCLC randomized to receive either nivolumab monotherapy or nivolumab + ipilimumab combination therapy and found no significant difference in mortality rates between these groups.

- Note: Since FDA approval in October 2018, combination therapy with pembrolizumab + chemotherapy has gradually replaced chemotherapy as first-line systemic treatment for patients with squamous aNSCLC. The NCT02785952 study was performed between 2016-2018.

**Gaps in knowledge:** While the proposed research addresses the innovative use of synthetic data in constructing ECAs for single-arm trials, several gaps in knowledge exist:

1. What metrics should be used to evaluate utility and privacy risks of synthetic ECA compared to the original,
2. What validation strategies should be used to ensure that the synthetic ECA captures inferential results compared to the original ECA
3. What best practices should be followed to ensure reliable synthetic ECA are generated.

The concept of using generative models to simulate data for “synthetic patients” to create a “virtual cohort” for the purposes of emulating a randomised trial has similarities to existing work in *in silico* clinical trials.

**What is the expected contribution of this study?** This study aims to answer the gaps in knowledge described above using a case study in mNSCLC, as well as describe limitations of this strategy and long-term implications for wider adoption.

## 6. Research question and objectives

**Table 2.** Primary and secondary research questions and objective.

A. Primary research question and objective
|  |  |
| --- | --- |
| **Objective:** | To compare the efficacy of nivolumab + ipilimumab combination therapy (“experimental arm”) versus nivolumab monotherapy (“control arm”) in patients with metastatic non-small cell lung cancer (mNSCLC) using real-world data (“original ECA”) and synthetic data derived from the same (“synthetic ECA”), with the aim of validating synthetic data for use in external control arm analysis. |
| **Hypothesis:** | This is not a hypothesis-driven study. However, we anticipate that synthetic ECA will be able to recapitulate an original ECA on different benchmarking metrics. |
| **Population (*mention key inclusion-exclusion criteria*):** | The following are eligibility criteria for NCT02785952.<br>• Age 18 years or older at index date<br>• Diagnosed with pathologically proven stage IV or recurrent squamous NSCLC<br>• No mixed histologies<br>• No other previous untreated malignancies<br>• Progression on one previous treatment with platinum-based chemotherapy<br>• No EGFR mutation or ALK fusion<br>• Sufficient tumour tissue for biomarker analysis<br>• ECOG score of 0 or 1<br>• No prior treatment with anti-PD-(L)1/2, CTLA-4 or any immune checkpoint inhibitor<br>• No active, known or suspected autoimmune diseases except some specific exceptions<br>• No known allergy or reaction to nivolumab and ipilimumab formulations<br>• No prior systemic treatment with corticosteroids or immunosuppressants within last 14 days<br>• No hepatitis B or C antibodies or infection, or HIV or AIDS<br>• No interstitial lung disease<br>• No grade III/IV cardiac disease or myocardial infarction within past 6 months<br>The target trial specification is provided below. |
| **Exposure:** | Nivolumab + ipilimumab combination therapy |
| **Comparator:** | Nivolumab monotherapy |
| **Outcome:** | Overall survival |
| **Time (*when follow up begins and ends*):** | Follow-up begins at the date of initiation of the index therapy ("index date") and ends at month 40 after index date or death, whichever is earlier. |
| **Setting:** | Stage IV squamous NSCLC |
| **Main measure of effect:** | Hazard ratio and restricted mean survival time over 40 months from index date |

| Protocol element | Target trial | Emulation (tentative) |
| --- | --- | --- |
| **Eligibility criteria** | • Age 18 years or older at index date<br>• Diagnosed with pathologically proven stage IV or recurrent squamous NSCLC<br>• No mixed histologies<br>• No other previous untreated malignancies<br>• Progression on one previous treatment with platinum-based chemotherapy<br>• No EGFR mutation or ALK fusion<br>• Sufficient tumour tissue for biomarker analysis<br>• ECOG score of 0 or 1<br>• No prior treatment with anti-PD-(L)1/2, CTLA-4 or any immune checkpoint inhibitor<br>• No active, known or suspected autoimmune diseases except some specific exceptions<br>• No known allergy or reaction to nivolumab and ipilimumab formulations<br>• No prior systemic treatment with corticosteroids or immunosuppressants within last 14 days<br>• No hepatitis B or C antibodies or infection, or HIV or AIDS<br>• No interstitial lung disease<br>• No grade III/IV cardiac disease or myocardial infarction within past 6 months | Same as target trial, except those in red, which are potentially unavailable or difficult to operationalize in real world datasets:<br>• Age 18 years or older at index date<br>• Diagnosed with pathologically proven stage IV squamous NSCLC<br>• No mixed histologies<br>• No other previous untreated malignancies<br>• Progression on one previous treatment with platinum-based chemotherapy<br>• No EGFR mutation or ALK fusion<br>• Sufficient tumour tissue for biomarker analysis<br>• ECOG score of 0 or 1 [not generally available for all patients; patients who meet other eligibility criteria and receive systemic treatment of cancer may be inferred to have an eligible performance status]<br>• No prior treatment with anti-PD-(L)1/2, CTLA-4 or any immune checkpoint inhibitor<br>• No active, known or suspected autoimmune diseases except some specific exceptions<br>• No known allergy or reaction to nivolumab and ipilimumab formulations<br>• No prior systemic treatment with corticosteroids or immunosuppressants within last 14 days<br>• No hepatitis B or C antibodies or infection, or HIV or AIDS<br>• No interstitial lung disease<br>• No grade III/IV cardiac disease or myocardial infarction within past 6 months |
| **Treatment regimens** | Initiation of (assignment to) one of | Same as target trial. However, information on whether the flat dose or the weight-adjusted dosage for nivolumab was prescribed would not generally be available. Therefore, |
|  | • Nivolumab administered intravenously at a dose of 3 mg/kg every 14 days ("control arm")<br>• Nivolumab administered intravenously at a dose of 3 mg/kg every 14 days + ipilimumab given at 1 mg/kg on day 1 of every third cycle ("experimental arm") | nivolumab initiation at any dosage compatible with observed data will be used. Notably, the flat dose and the weight-adjusted dose regimens show the same safety and efficacy profiles in prior studies (Bei et al. 2020) |
| **Treatment assignment** | Patients were randomly assigned to the control or experimental arm at baseline | Treatment assignment is assumed random conditional on measured risk factors, potentially including age at baseline, sex, ECOG score, presence of liver and bone metastases and index line of therapy |
| **Outcome** | All-cause mortality | Same as target trial |
| **Follow-up** | Begins at treatment assignment and ends at month 40, study discontinuation or death, whichever occurs first | Same as target trial |
| **Causal contrast** | Intention to treat effect on the absolute and relative scales, namely restricted mean survival time and hazard ratio | Same as target trial |
| **Statistical analysis** | Kaplan-Meier curves will be reported and restricted mean survival will be calculated using these curves. Hazard ratio will be estimated using Cox proportional hazard model. | Kaplan-Meier curves and hazard ratios will be estimated using models weighted by the inverse probability of treatment assignment. Inverse weights will be estimated using a logistic regression model with treatment assignment indicator as a function of baseline confounders. Point estimates and 95% confidence intervals based on robust standard errors to account for weighting will be documented. |

The analysis will be replicated with no change using the original and synthetic ECAs.

## 7. Research methods

### 7.1. Study design

**Research design (e.g. cohort, case-control, etc.):** External control arm study (non-randomized comparison of trial and real-world patients)

**Rationale for study design choice:** For assessing the feasibility of using synthetic data for ECA analysis, an ECA design is most appropriate.

### 7.2. Study design diagram

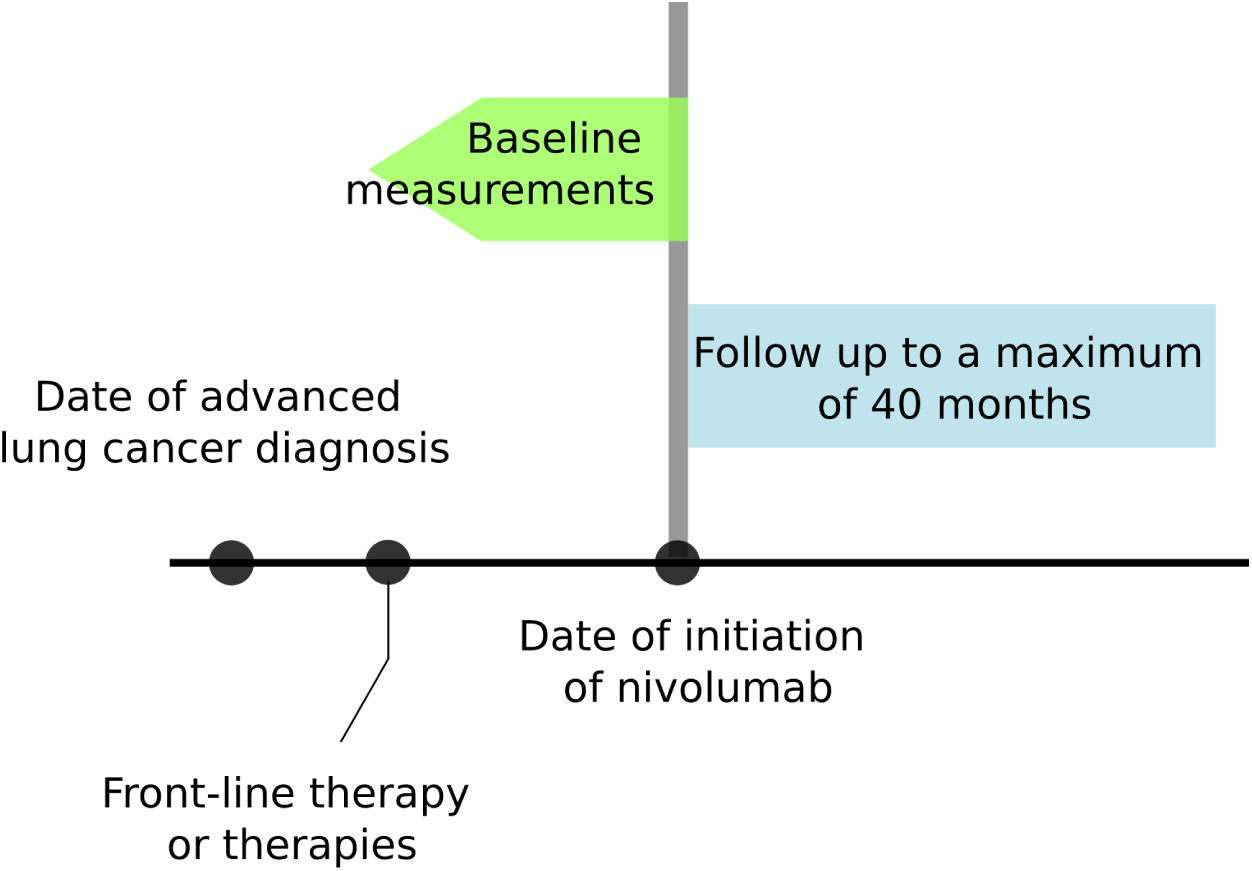

### 7.3. Setting

#### 7.3.1 Context and rationale for definition of time 0 (and other primary time anchors) for entry to the study population

The index date is the date of treatment assignment in the Lung-MAP study NCT02785952. The ECA analysis aims to emulate the same.

**Table 3.** Operational Definition of Time 0 (index date) and other primary time anchors.

| Study population name(s) | Time Anchor Description (e.g. time 0) | Number of entries | Type of entry |
| --- | --- | --- | --- |
| NCT02785952 | Date of randomization | 1 | Incident use |

#### 7.3.2 Context and rationale for study eligibility criteria

Eligibility criteria for the target trial are identical to those from NCT02785952.

**Table 4.**
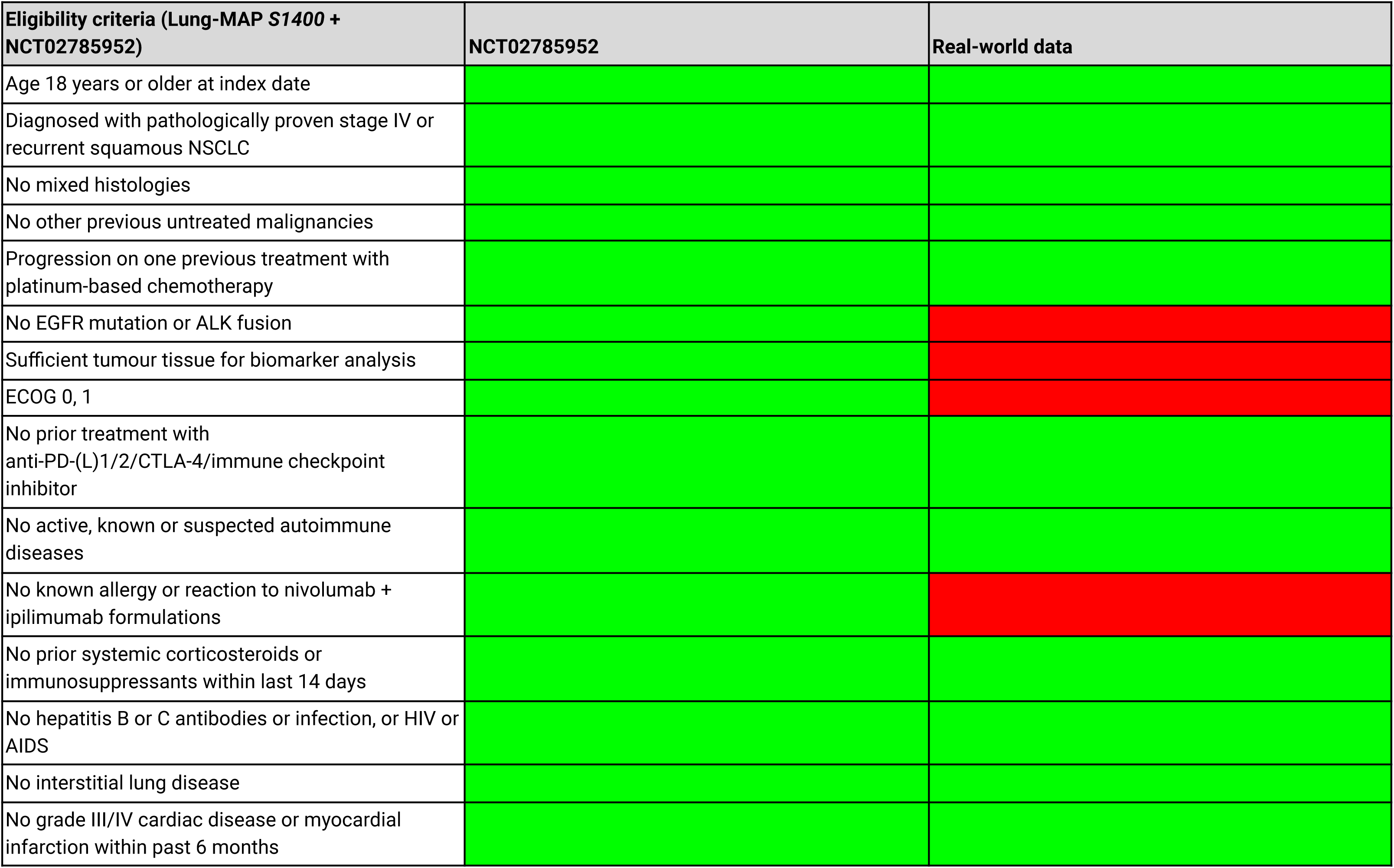
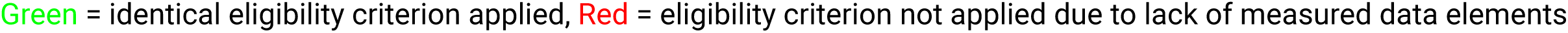
Operational Definitions of Inclusion/Exclusion Criteria.

### 5.4. Variables

#### 7.4.1 Context and rationale for exposure(s) of interest

The treatment groups of interest are the same as those in NCT02785952. For real-world studies, we make the concession that any dose and frequency compatible with observed data is permitted, even if not reported.

Treatment regimens in NCT02785952:

● Nivolumab administered intravenously at a dose of 3 mg/kg every 14 days
● Nivolumab administered intravenously at a dose of 3 mg/kg every 14 days + ipilimumab given at 1 mg/kg on day 1 of every third cycle

**Algorithm to define duration of exposure effect:**

n/a

#### 7.4.2 Context and rationale for outcome(s) of interest

Overall survival defined as time from index date to death from any cause is the primary and sole outcome of interest. Overall survival is the most important clinical outcome in aNSCLC.

**Table 7.**
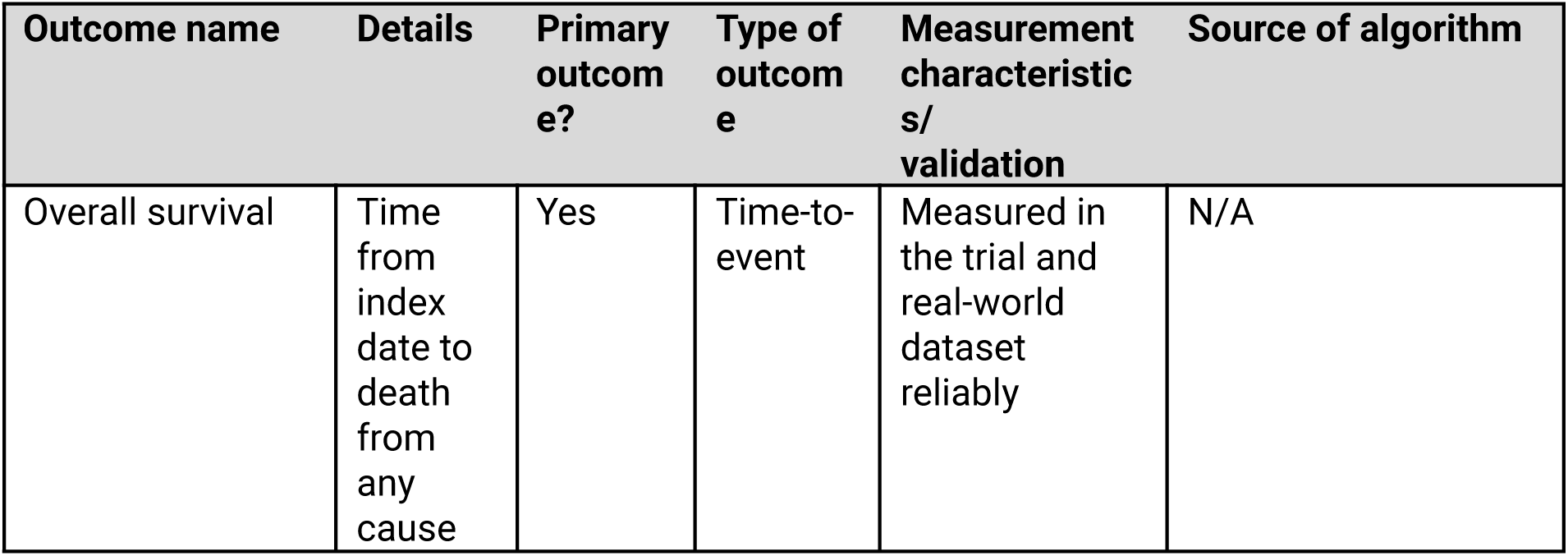
Operational Definitions of Outcome.

#### 7.4.3 Context and rationale for follow up

The maximum length of follow-up in the NCT02785952 data cut available to us is approximately 40 months.Click here to enter text.

**Table 8.**
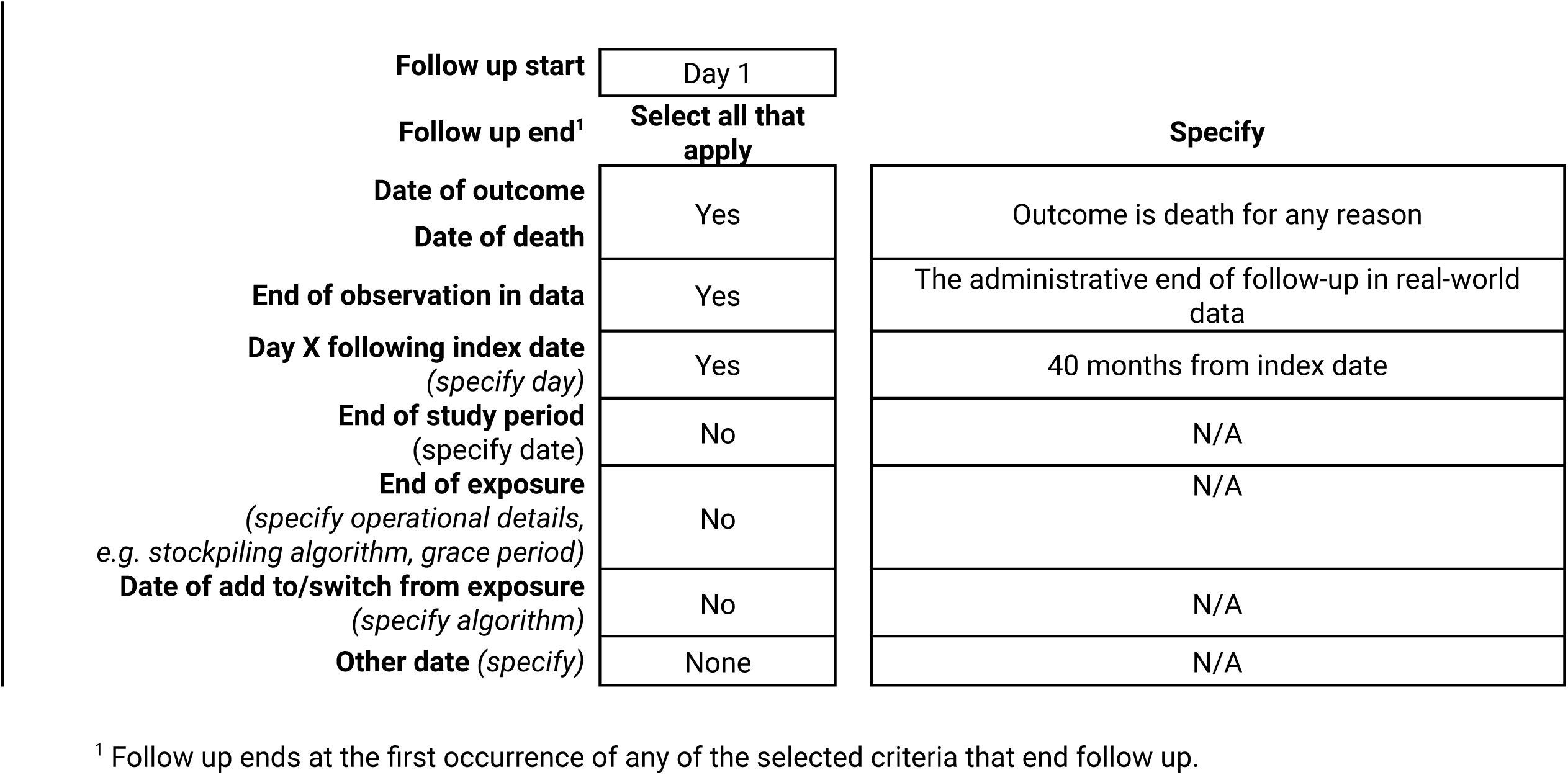
Operational Definitions of Follow Up.

#### 7.4.4 Context and rationale for covariates (confounding variables and effect modifiers, e.g. risk factors, comorbidities, comedications)

An assumption for unbiased estimation of the treatment effect is adjustment for all confounders of the treatment effect. However, not all risk factors for survival are measured across the data sources. Furthermore, it is impossible to adjust for all risk factors even if they were measured. We only adjust for measured confounding variables in this study.

**Table 9.** Operational Definitions of Covariates. The following is a tentative list of measured risk factors for overall survival we will attempt to adjust for. Variables marked with an asterisk are missing for the majority of patients in the real-world database.

| Characteristic | Details | Type of variable | Assessment window |
| --- | --- | --- | --- |
| Age | Age (in years) at randomization | Continuous | Screening variables were assessed within 14 days of registration in NCT02785952 |
| Sex | Sex | Dichotomous (male or female) |  |
| ECOG score* | ECOG performance score (0-1) | Dichotomous |  |
| Liver metastases | Presence of liver metastases at baseline | Dichotomous (present or absent) |  |
| Brain metastases | Presence of brain metastases at baseline | Dichotomous (present or absent) |  |
| Smoking history* | History of smoking | Dichotomous (ever or never) |  |
| Index line | Index line of therapy | Dichotomous (1 or 2+) |  |

### 7.5. Data analysis

#### 7.5.1 Context and rationale for analysis plan

The inferential data analysis plan is described conceptually, and the context or rationale for the choices are provided in this section.

**Table 10.**
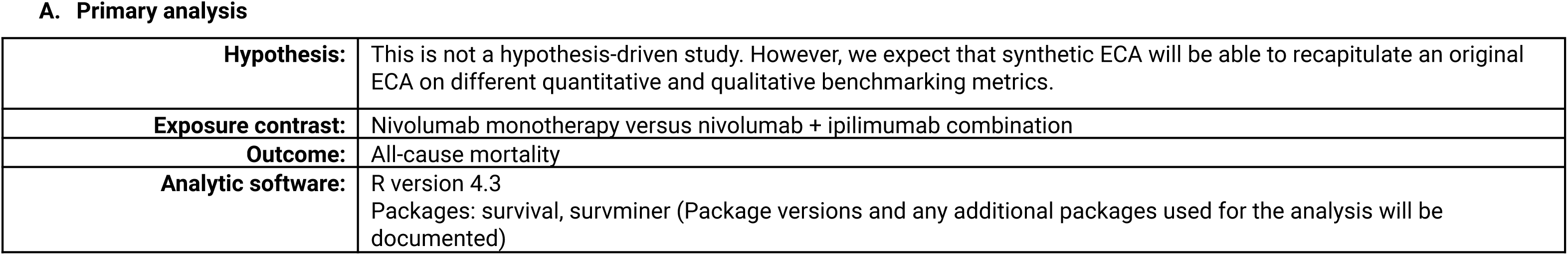
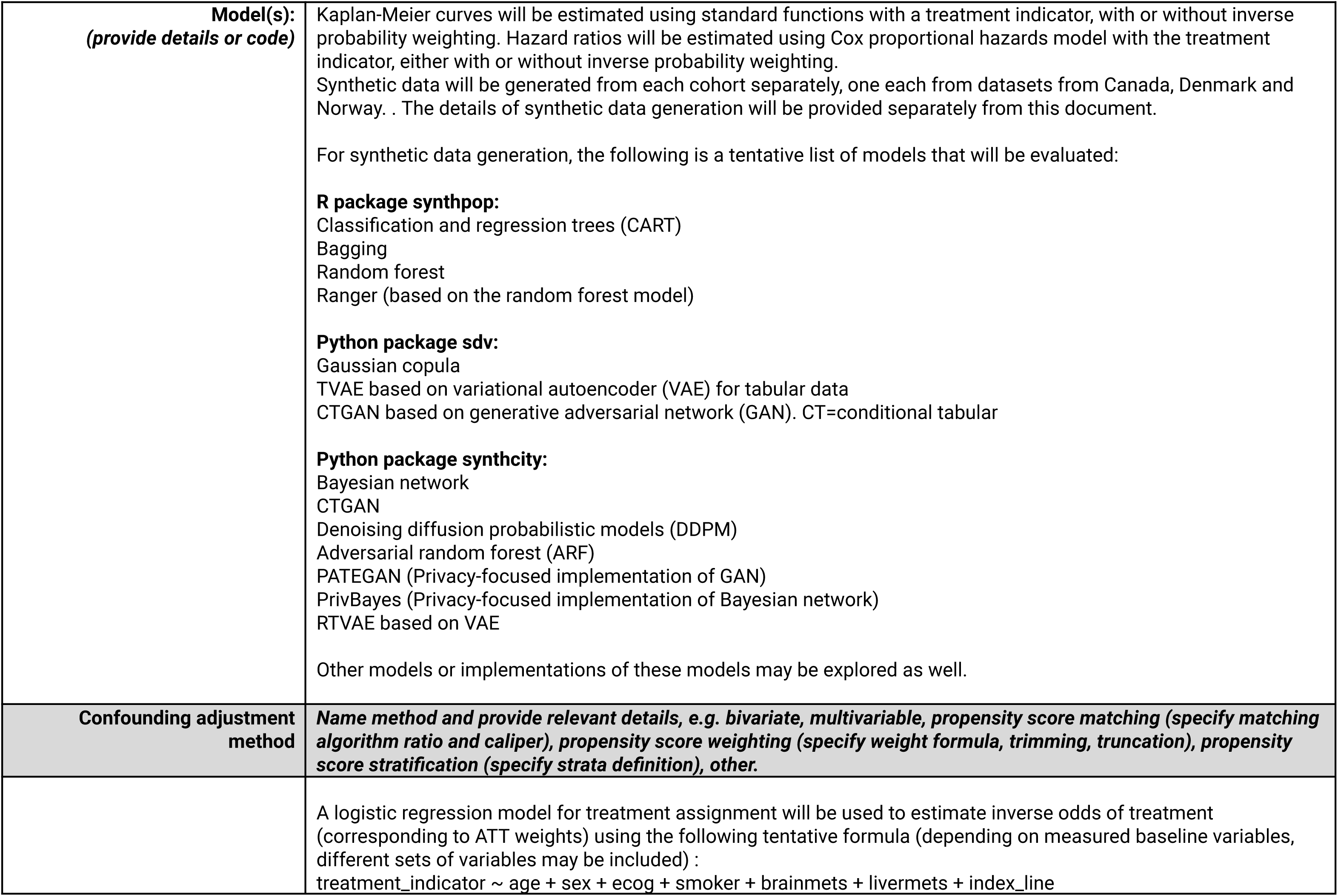
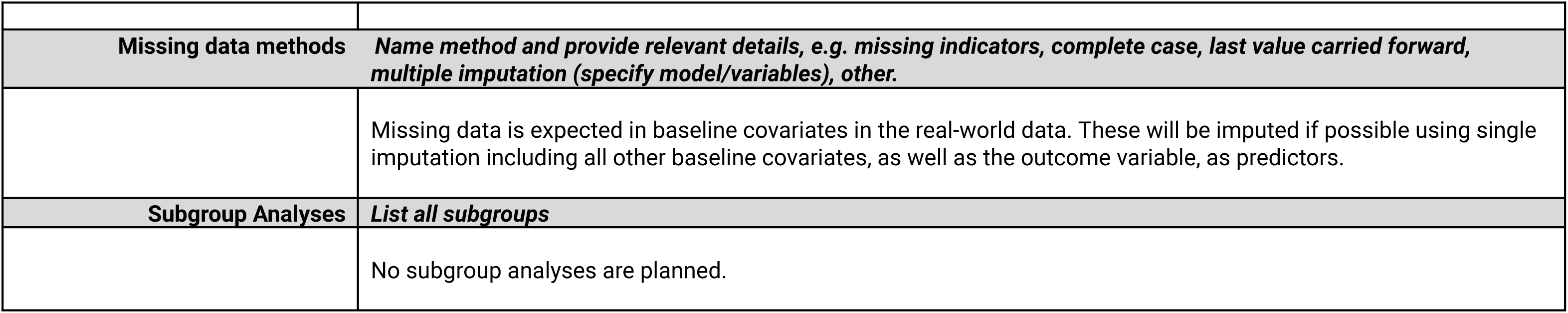
Primary, secondary, and subgroup analysis specification.

**Table 11.**
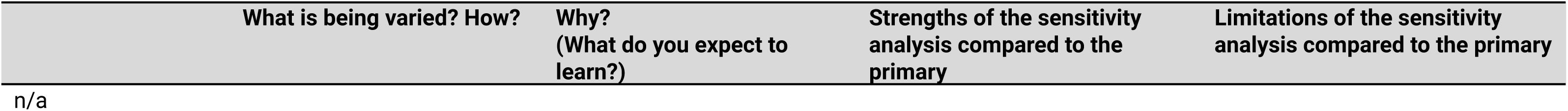
Sensitivity analyses – rationale, strengths and limitations. No sensitivity analyses are planned.

### 7.6. Data sources

#### 7.6.1 Context and rationale for data sources

**Reason for selection:** Selection of data sources was based on availability of individual-level data. NCT02785952 was selected from Project Data Sphere. The criteria for selecting a suitable clinical trial dataset for this study were as follows: (i) a lung cancer indication, (ii) a randomized trial with both treatment arms available, (iii) sample size >100 and (iv) testing a non-chemotherapy regimen of clinical importance that is approved for use and commonly administered. NCT02785952 was the only trial on Project Data Sphere that fit these criteria. aNSCLC was chosen because the study authors have substantial experience in this disease setting, and because it is a common indication for drug development, and therefore for regulatory approvals and health technology assessments.

Subsequently, three real-world data sources were identified:

● Administrative cancer data from the province of Alberta, Canada
● Danish national cancer registry
● Norwegian national cancer register

**Strengths of data source(s):** Patient-level data is available for important risk factors and overall survival.

**Limitations of data source(s):** The original ADaM/SDTM dataset is not available, and therefore we only have derived variables in some cases for the trial. No longitudinal data on risk factors or subsequent therapies is available. Some variables, notably ECOG scores, are not available in the real-world data.

**Data source provenance/curation:** Not available for the trial dataset. However, the distribution of baseline characteristics and Kaplan-Meier estimates match those from the trial publication (not shown here). Both the Canadian and Nordic databases collect cancer data at a national population level and are well-documented and have previously been used for epidemiologic and comparative analyses.

**Table 12.** Metadata about data sources and software.

|  | **Trial data** | **Real-world data (Canada)** | **Real-world data (Denmark)** | **Real-world data (Norway)** |
| --- | --- | --- | --- | --- |
| **Data Source(s):** | NCT02785952 | Alberta Health | Danish cancer register | Norwegian cancer registry |
| **Study Period:** | 2015-12-29 to 2019-12-19 based on https://www.clinicaltrials.gov/study/NCT02785952. | Although data from a broader study period is available across all three real-world databases (e.g, 2008-2022 for Nordic countries), only data from the trial study period ranging from 2015 to 2019 will be used. |  |  |
| **Eligible Cohort Entry Period:** | Not known. The data available does not include this information. | See target trial specification. |  |  |
| **Data Version (or date of last update):** | Not known | TBD | TBD | TBD |
| **Data sampling/extracti on criteria:** | n/a | See target trial specification | See target trial specification | See target trial specification |
| **Type(s) of data:** | Patient-level data from a randomized clinical trial | Population-based data from the Canadian province of Alberta. Patients with advanced lung cancer identified from the Alberta Cancer Registry, which captures information on more than 99% of cancers diagnosed in the province | Population-based registry from patients with incident cancer diagnosis in Denmark | Population-based registry from patients with incident cancer diagnosis in Norway |
| **Data linkage:** | n/a |  |  |  |
| **Conversion to CDM\*:** | None | None | None | None |
| **Software for data management:** | None |  |  |  |

\*CDM = Common Data Model

### 7.7. Data management

Patient-level data from NCT02785952 has been provided by the trial sponsor in a deidentified format. A single copy of this dataset will be stored on a local password-protected computer. Programming code will be stored and backed up in a private and secured cloud repository.

### 7.8. Quality control

For NCT02785952, we have a limited dataset provided by the trial sponsor. Data from Alberta, Canada and Nordic countries are both validated and analyzed by an expert deeply familiar with the data and its limitations. Although double programming will not be performed, steps will be taken to ensure that there are no programming errors for data processing and analysis that can affect the accuracy of the results, for example, through sanity checks and visualization of intermediate results/outputs in the analysis.

### 7.9. Study size and feasibility

Because this is an exploratory study, sample size calculations were not formally performed. The primary objective of this study is to compare results between original ECA and synthetic ECA used for ECA analysis, rather than producing unbiased estimates of the treatment effect (which are available from the trial and known). Therefore, the study should be feasible regardless of sample size or adequacy of confounding control. However, sensitivity analyses will be performed for unmeasured confounding.

**Table 13.** Power and sample size. Not applicable. This is an exploratory study and no formal power calculations were performed.

## 8. Limitation of the methods

The following is a discussion of potential limitations of the study design, and analytic methods, including issues relating to confounding, bias, generalisability, and random error:

1. Random error – NCT02785952 is a relatively small trial (125 + 127 patients) and therefore the results may have low precision. This limitation exists for any study with small samples, and is a common concern in external control arm studies. Therefore, it may be difficult to disentangle a small difference in results between the original ECA and synthetic ECA if it exists.
2. Inability to emulate the target trial perfectly – Due to lack of data availability, there are limitations for perfectly emulating eligibility criteria from the trial. There is also the possibility that there are other unknown differences in variable recording and derivation across the two datasets. However, the primary objective of this study is to compare results between original ECA and synthetic ECA used for ECA analysis, rather than producing unbiased estimates of the treatment effect, and therefore this limitation may be discussed by comparing ECA results with those from the randomized trial.
3. Generalizability of the results – Our results may not generalize to other settings or other datasets. We will not assert that they do. Instead, we will use this case study to describe best practices for future benchmarking studies or applications using synthetic data for ECA analyses.

## 9. Protection of human subjects

n/a. This study uses deidentified data.

## 10. Reporting of adverse events

n/a

## Data Availability

All data produced in the present study are available upon request to the relevant coauthors.

## 12. Appendices

n/a

## References

1. Sabbula BR, Gasalberti DP, Anjum F. Squamous cell lung cancer. InStatPearls [Internet] 2023 Sep 4. StatPearls Publishing.

2. Gettinger SN, Redman MW, Bazhenova L, Hirsch FR, Mack PC, Schwartz LH, Bradley JD, Stinchcombe TE, Leighl NB, Ramalingam SS, Tavernier SS. Nivolumab plus ipilimumab vs nivolumab for previously treated patients with stage IV squamous cell lung cancer: the lung-MAP S1400I phase 3 randomized clinical trial. JAMA oncology. 2021 Sep 1;7(9):1368–77.

3. Thorlund K, Dron L, Park JJ, Mills EJ. Synthetic and external controls in clinical trials–a primer for researchers. Clinical epidemiology. 2020 May 8:457–67.

4. Arora A. Synthetic data: the future of open-access health-care datasets?. The Lancet. 2023 Mar 25;401(10381):997.

5. Bei D, Osawa M, Uemura S, Ohno T, Gobburu J, Roy A, Hasegawa M. Benefit-risk assessment of nivolumab 240 mg flat dose relative to 3 mg/kg Q2W regimen in Japanese patients with advanced cancers. Cancer Science. 2020 Feb;111(2):528–35.

6. Pathmanathan P, Aycock K, Badal A, Bighamian R, Bodner J, Craven BA, Niederer S. Credibility assessment of in silico clinical trials for medical devices. PLOS Computational Biology. 2024 Aug 8;20(8):e1012289.

